# Differential expression of fibroblast activation protein-alpha and lysyl oxidase in subtypes of ameloblastoma

**DOI:** 10.1101/2023.03.10.23286752

**Authors:** Tim Vangansewinkel, Ronald Driesen, Jimoh Olubanwo Agbaje, Pascal Gervois, Akinyele Olumuyiwa Adisa, Adeola Adenike Olusanya, Juwon Tunde Arotiba, Esther Wolfs, Ivo Lambrichts, Constantinus Politis

## Abstract

Ameloblastomas are benign, mostly intra-osseous, tumours originating from ectodermal odontogenic epithelium and display extracellular matrix remodelling. We determined whether fibroblast activation protein-alpha (FAP-α), a collagenolytic enzyme and lysyl oxidase (LOX), a collagen cross-linker were differentially regulated in ameloblastoma subtypes. Masson’s trichrome staining and immunohistochemistry were performed on human samples from gross mandibular or maxillary ameloblastoma. Co-localization studies in tumorigenic tissue of follicular, plexiform and desmoplastic ameloblastoma showed absence of the mesenchymal cell marker vimentin but high epithelial cell adhesion molecule (EPCAM) expression, an epithelial marker. Strong FAP-α staining was also noted, however, the stellate reticulum of plexiform ameloblastoma contained a higher number of FAP-α positive cells than the follicular subtype. Weak LOX staining was demonstrated in tumorigenic tissue of the follicular subtype but profound reactivity was observed in stromal plexiform and tumorigenic desmoplastic tissue. The desmoplastic staining profile for FAP-α and LOX shared resemblance with the unicystic subtypes. Tumorigenic outgrowths in desmoplastic ameloblastoma were associated with vimentin positive and EPCAM negative cancer-associated fibroblasts suggesting a role in tumour invasion. In conclusion, FAP-α expression in ameloblastoma seems to be involved in tumour growth and invasion whereas LOX expression is differentially regulated in ameloblastoma subtypes offering a new perspective for understanding specific growth patterns of ameloblastoma subtypes.

## Introduction

Ameloblastomas are slow growing benign intra-osseous tumours which originate from the ectodermal odontogenic epithelium. They are mainly located in the lower jaw and more specifically in the posterior mandible, except for the desmoplastic ameloblastoma which has a predilection for the anterior region. The total incidence rate of ameloblastomas worldwide is limited to 0.92 per million person-years ^1^ but varies geographically. Prevalence of benign ameloblastoma is noted to be the highest in Asian and African countries^2^ particular in Nigeria^3^. Sixty percent of all Nigerian patients with ameloblastoma have a young age between 18 and 40 years old with the posterior mandible as the most common site of tumour growth^4^. Ameloblastomas range from entirely solid to variably cystic. The most prevailing histopathological solid pattern is the follicular, followed by the plexiform ameloblastoma. Other histopathological types include acanthomatous, granular, basaloid, clear-cell, hemangiomatous, hybrid, keratoameloblastoma and the desmoplastic type. The peripheral ameloblastoma is only found in oral mucosa and adjacent soft tissues^1,5,6^. The different histopathological patterns are observed separately or combined within the tumour. Unicystic ameloblastoma is also located intra-osseous, appears as a cystic cavity and is classified based on the location of epithelial tumorigenic growth i.e. luminal (type I), intraluminal (type II) and mural (type III)^6^. Desmoplastic ameloblastoma is the rarest variant of the solid ameloblastoma and is composed of connective tissue intermingled with small irregular tumorigenic islands^7^. A histological characteristic of tumorigenic tissue in all subtypes is the presence of ameloblast-like cells at the epithelial border and the adjacent growth of stellate reticulum, depending on the variant. Therapy is based on surgical resection however lifetime follow-up is required to limit tumour recurrence. The aetiology of ameloblastomas has a multifactorial origin however at the gene expression level, a mutation in the BRAFV600E gene is found in the majority of all subtypes^8,9^. Disturbed regulation of signalling pathways such as sonic hedgehog (SHH), Wingless/integrase 1 (WNT) type 5A, and mitogen-activated protein kinase (MAPK) have been reported to induce oncogenic activation of odontogenic epithelium^1,10^. Recent studies postulate that ameloblastoma growth and invasion is not only determined by genomic alterations but coincides with a direct interaction with cancer-associated fibroblasts (CAF) in the adjacent stromal tissue^11^. CAF share structural and functional similarities with myofibroblast^12^, secrete proteolytic enzymes which remodel the extracellular matrix (ECM), and aid in the directional growth and migration of tumour cells^13^. More specifically, fibroblast activation protein-alpha (FAP-α) has been identified in CAF from epithelial carcinomas and its expression is associated with enhanced tumour growth^14,15^. FAP-α is part of the family of surface serine proteases and displays collagenase activity^15^. FAP-α is mainly expressed during embryogenesis and is downregulated when reaching adolescence^16^. FAP-α re-expression is driven by TGF-β1 and interleukin-1β^17^, and is observed in pathologies associated with ECM remodelling, fibroblast differentiation^18^, and in the process of epithelial to mesenchymal cell transition (EMT)^19^. Rapid progression in tumour development has been linked to a gradual increase in ECM stiffness which is associated with enhanced invasion capacity, proliferation, and lysyl oxidase (LOX) expression^20^. LOX belongs to the same stromal gene expression program as FAP-α^21^, is produced by CAF and aids in stabilizing the ECM by cross-linking collagen type I and elastin. In addition, LOX expression enhances tumour cell migration, promotes invasion and stimulates EMT^22^. In this study, we hypothesized that the specific growing patterns of ameloblastoma subtypes are associated with differential regulation of FAP-α and LOX expression within the stromal and tumorigenic compartments of the tissue. We sought to determine whether FAP-α and LOX expression levels are linked to ECM remodelling, invasion and collagen type I crosslinking in subtypes of ameloblastoma.

## Methods

### Ethical statement, patient samples and ameloblastoma subtyping

This study was approved by the Medical Ethical Committee in Nigeria (ADM/DCST/HREC/APP/753). The research was conducted in accordance with the 1975 Helsinki Declaration and its later amendments. Fourteen consenting patients (nine males and five females; aged between 15 – 44 years old) were included in this study presenting gross ameloblastoma of the mandible or maxilla. Study groups (Table 1) were created based on the radiographic appearance and/or histological analysis i.e. unicystic, solid follicular (+ or - acanthomathous), solid plexiform (+ or - acanthomathous) and desmoplastic (follicular and plexiform). Ameloblastoma samples from complete tumour resection were fixed in 4% paraformaldehyde and embedded in paraffin. Serial sections (5 μm) were deparaffinised, rehydrated prior to Masson’s Trichrome staining and mounted in DPX (Sigma-Aldrich). The sections were histologically examined by a pathologist (Dr. Adisa AO) to determine the ameloblastoma subtype.

**Table 1:** Classification of the ameloblastoma samples.

| **Type of ameloblastoma based on radiography** | **Histological classification** | **Gender** | **Localization** |
| --- | --- | --- | --- |
| Unicystic | Undefined | Female | Mandible |
| Unicystic | Undefined | Female | Mandible |
| Unicystic | Undefined | Male | Mandible |
| Solid | Follicular | Male | Mandible |
| Solid | Follicular acanthomatous | Female | Mandible |
| Solid | Follicular acanthomatous | Male | Maxilla |
| Solid | Follicular acanthomatous | Female | Mandible |
| Solid | Plexiform | Female | Maxilla |
| Solid | Plexiform | Male | Mandible |
| Solid | Plexiform acanthomatous | Male | Mandible |
| Desmoplastic | Follicular | Male | Mandible |
| Desmoplastic | Plexiform | Male | Mandible |
| Desmoplastic | Follicular | Male | Mandible |
| Desmoplastic | Follicular | Male | Mandible |

### Second harmonic generation and confocal microscopy

Second harmonic generation (SHG) label-free imaging was performed using a Zeiss LSM 880 (Carl Zeiss, Jena, Germany) mounted on an Axio Observer and equipped with a 20x objective (Plan-Apochromat 20x/0.8, Carl Zeiss). A pixel size of 0.69 mm was used with an image resolution of 1024 by 1024 and a pixel dwell time of 8.19 ms. Full tissue sections were imaged by means of tile scans with 10% overlap to enable the stitching of the recorded tiles. Excitation was provided by a femtosecond pulsed laser (MaiTai DeepSee, Spectra-Physics, CA, United States) tuned to a central wavelength of 810 nm. This laser was directed to the sample using a 760 nm short pass dichroic mirror. The SHG signal and autofluorescence were collected in backward non-descanned mode using a 760 nm short pass dichroic mirror. A BP 350-690 bandpass filter was used to block any scattering infrared light. Finally, a 425 nm dichroic mirror separated SHG from autofluorescence which were subsequently simultaneously recorded via, respectively, a 400–410 nm bandpass filter and a 450–650 nm band pass filter, by means of GaAsP detectors (BIG2, Carl Zeiss).

### Immunohistochemistry

Deparaffinised tissue sections were treated with heated citrate buffer (Dako, Glostrup, Denmark) to induce antigen retrieval followed by cooling down. To prevent background staining, sections were treated with peroxidase and protein block (Dako, Glostrup, Denmark). For DAB immunostaining, sections were incubated with primary antibodies (Abcam, Cambridge, UK) against FAP-α (1:200, Ab53066, rabbit anti-human), vimentin (1:100, Ab8069, mouse anti-human), EPCAM (1:100, Ab71916, rabbit anti-human) and LOX (1:200, Ab174316, rabbit anti-human) diluted in PBS for 1 hour at room temperature followed by washes with PBS. As negative control, the primary antibody was omitted from a section. Peroxidase-conjugated secondary antibodies (goat anti-rabbit or goat anti-mouse) diluted in PBS were applied for 45’ at room temperature followed by washes in PBS. The chromogenic substrate 3,3’-Diaminobenzidin (DAB) was used to visualize the protein of interest (DAB kit, Dako, Glostrup, Denmark). Sections were counterstained with Mayer’s haematoxylin and mounted using DPX mounting medium. Immune-reactivity was determined using a photomicroscope equipped with an automated camera (Nikon Eclipse 80i, Nikon Co., Japan). Quantification of FAP-α staining was performed in serial sections using the cell counter tool in ImageJ (National Institutes of Health, http://rsb.info.nih.gov/ij/).

### Statistical analysis

All statistical analyses were performed using GraphPad Prism 5.01 software (GraphPad Software, Inc.). Data sets were analyzed for normal distribution using the D’Agostino-Pearson normality test. Statistical analysis was performed using one-way ANOVA with Bonferroni post-hoc test. The analyst was blinded by coding of the images. Data were presented as mean + standard error of the mean (SEM). Differences were considered statistically significant when p < 0.05.

## Results

### Collagen type I distribution and organization in subtypes of ameloblastoma

SHG microscopy, a label-free imaging method ^23^, revealed the presence of tumorigenic tissue in follicular ameloblastoma biopsies which was completely devoid of collagen type I. The tumorigenic tissue is surrounded by densely packed connective tissue which is composed of a high concentration of collagen type I (Fig. 1A, B). The tissue from plexiform ameloblastoma showed a markedly weaker signal of collagen type I compared to follicular ameloblastoma and is visible as thinner bundles of cross-linked collagen type I (Fig. 1C, D). The desmoplastic ameloblastoma tissue consists mainly of connective tissue with small strands of tumorigenic tissue enclosed by collagen type I (Fig. 1E, F).

**Figure 1:**
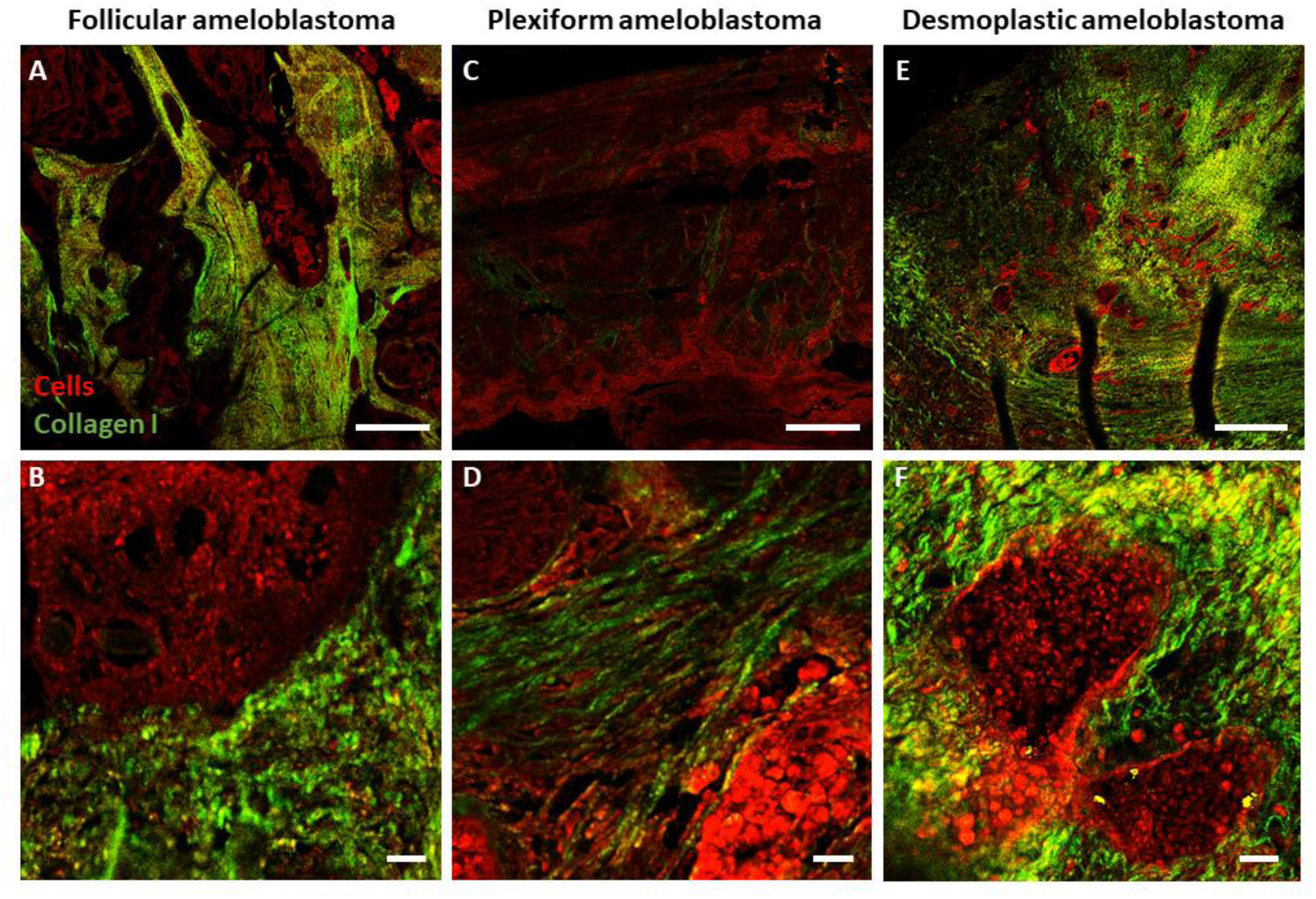
Visualization of collagen type I in subtypes of ameloblastoma. The label-free imaging technique second harmonic generation (SHG) confocal microscopy was used to detect collagen type I (in green) in follicular (A, B), plexiform (C, D) and desmoplastic (E, F) ameloblastoma. The autofluorescence signal (in red) was used to visualize cells in the different ameloblastoma tissues. Scale bars represent 500 (A, C, E) and 20 (B, D, F) μm.

### Determination of FAP-α expression in follicular and plexiform ameloblastoma

Masson’s Trichrome staining of follicular ameloblastoma biopsies showed a dense collagen matrix enclosing the tumorigenic tissue (Figure 2A). The latter is composed of a thin epithelial border of ameloblast-like tumour cells and an inner stellate reticulum containing spindle-shaped tumour cells (Figure 2B). Collagen was completely devoid within the tumorigenic tissue. Co-localization studies in serial sections (Fig. 2C-J) revealed an almost complete absence of vimentin staining in the tumorigenic tissue (Fig. 2E, F, L) except for some occasional cells (Fig. 2L, arrow) and a high expression of the epithelial marker EPCAM in the ameloblast-like cells and stellate reticulum (Fig. 2G, H). The surrounding stroma demonstrated a strong vimentin staining (Fig. 2M, arrows) in the spindle-shaped CAF but these were negative for EPCAM (Fig. 2N). LOX staining within the same region showed a weak expression profile in the tumorigenic tissue (Fig. 2I, J). Masson’s Trichrome staining of plexiform ameloblastoma tissues revealed the ECM encompassing the large tumorigenic tissue (Fig. 3A, B). FAP-α positive cells were present in the ameloblast-like epithelial layer and stellate reticulum (Fig. 3C, D). As reported for the follicular type, the tumorigenic tissue of plexiform ameloblastoma was completely negative for vimentin (Fig 3E, F) but positive for EPCAM (Fig. 3G, H). LOX staining was mainly observed within the stroma (Fig. 3I; arrow) and delineated the transition between the connective and tumorigenic tissue (Fig. 3J, arrow).

**Figure 2:**
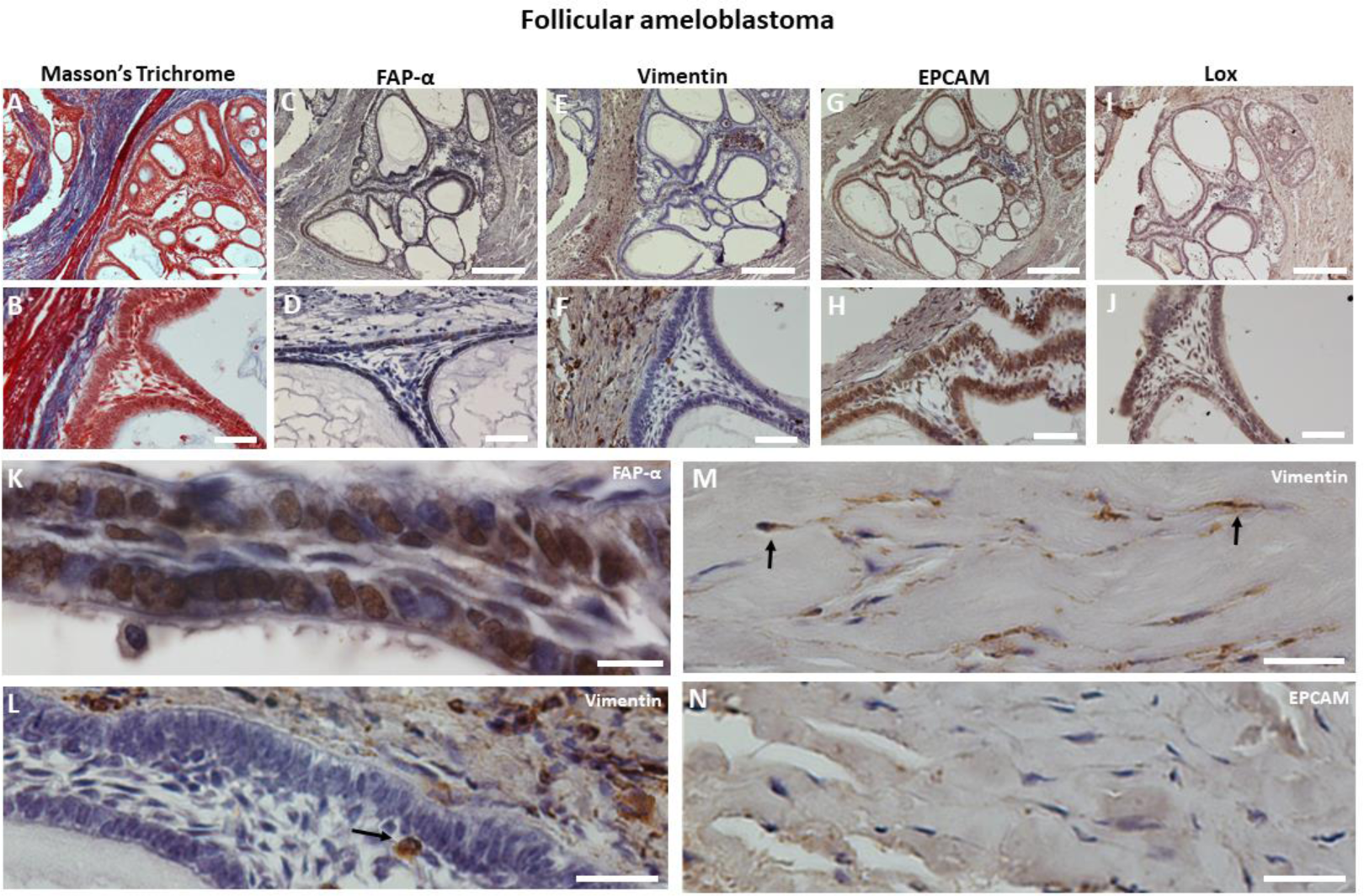
Identification of FAP-α and LOX expressing cells in follicular ameloblastoma. Histological evaluation of serial sections using Masson’s Trichrome staining (A, B; blue = collagen (stroma); red = tumorigenic tissue). Co-localization of FAP-α positive (C, D), vimentin negative (E, F), EPCAM positive (G, H) and LOX weak positive (I, J) expression in tumorigenic tissue using immunohistochemistry. Higher magnifications (K-N) indicate FAP-α positive (K) and vimentin (L) negative tumour cells. Note the presence of a vimentin positive cell (L, arrow). Spindle-shaped CAF in the stromal tissue are vimentin positive (M, arrows) but EPCAM (N) negative. Scale bars represent 25 (K), 100 (B, D, F, H, J), 50 (L, M, N), 500 (A, C, E, G, I) μm.

**Figure 3:**
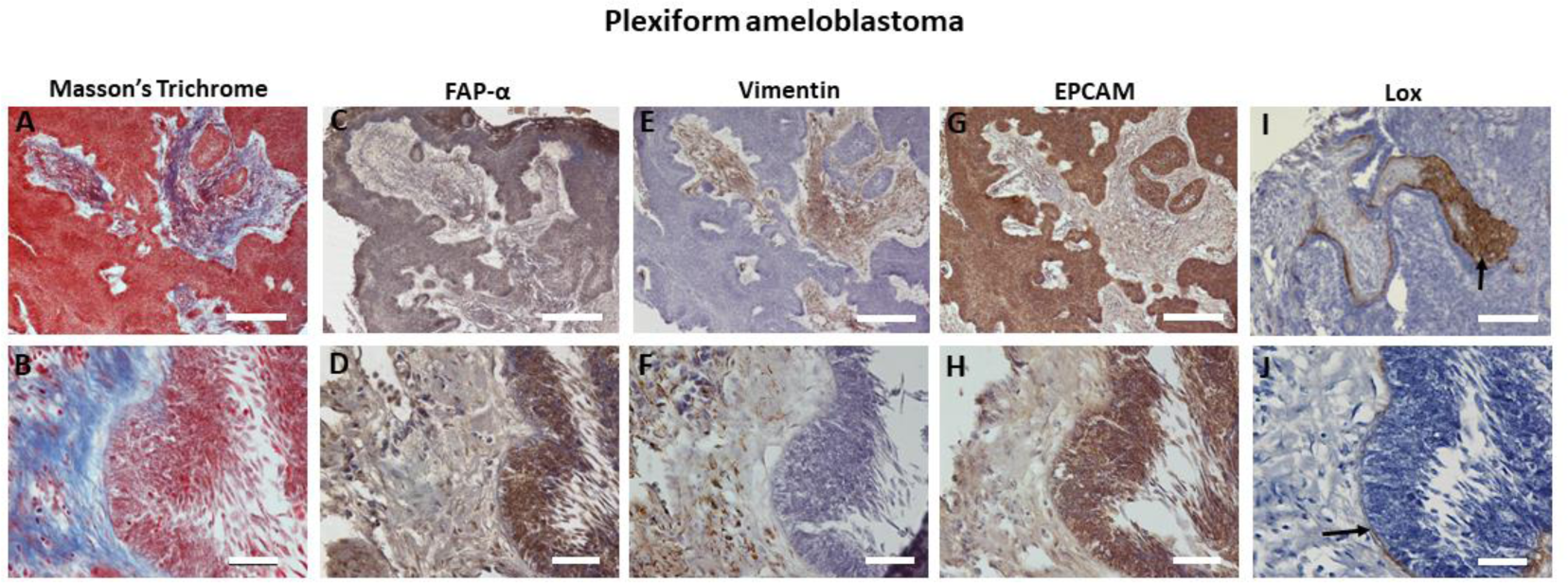
Identification of FAP-α and LOX expressing cells in plexiform ameloblastoma. Histological evaluation of serial sections using Masson’s Trichrome staining (A, B; blue = collagen (stroma); red = tumorigenic tissue). Co-localization of FAP-α positive (C, D), vimentin negative (E, F), EPCAM positive (G, H) and LOX negative (I, J) expression in tumorigenic tissue using immunohistochemistry. LOX expression is restricted to the stromal tissue (2I, arrow) and delineates the border between the stromal and tumorigenic tissue (J, arrow). Scale bars represent 100 (B, D, F, H, J), 500 (A, C, E, G, I) μm.

### FAP-α expression in desmoplastic ameloblastoma

Tissue of desmoplastic ameloblastoma is mainly composed of connective tissue with strands of tumorigenic tissue (Fig. 4A). Invasion into the stroma was detected by the presence of protrusions sprouting out from the tumorigenic mass. At the tips of these protrusions, a translucent cellular phenotype was observed (Fig. 4B, arrow) which was markedly different from the darker phenotype of the tissue of origin. These translucent cells seemed to be concentrated in small foci (Fig. 4C, arrow) with no complex tissue organization indicating a less differentiated phenotype. The translucent cells were engulfed and contained by highly cross-linked and dense collagen fibrils (Fig. 4D) when observed in a transverse orientation. Frequently, we detected tumorigenic islands with a distinct halo which showed a less dense collagen concentration (Fig. 4E). These tumorigenic cells displayed a darker and differentiated phenotype with features of ameloblastoma tumorigenic tissue i.e. an epithelial ameloblast-like cell lining and stellate reticulum. A longitudinal orientation confirmed the cross-linked collagen surrounding the translucent tumorigenic tissue (Fig. 4F) and at higher magnification spindle-shaped myofibroblast-like cells or CAF were detected interspersed between collagen fibres (Fig. 4F, arrows in G). FAP-α staining showed a weak staining in the less differentiated phenotype surrounded by dense connective tissue (Fig. 5A). However, strong staining was noted in the differentiated tumorigenic islands (Fig. 5B) and in small protrusions (Fig. 5C, arrows). Higher magnification identified FAP-α positive ameloblast-like cells in the epithelial lining (Fig. 5D) but was less pronounced in the stellate reticulum and stroma. We classified this tissue as tumorigenic due to the absence of vimentin staining (Fig. 5E) which was only observed in stromal cells and to the strong EPCAM staining (Fig. 5F). Occasionally, we detected vimentin positive cells within the epithelial border (Fig. 5E, insert). High LOX expression was demonstrated in tumorigenic tissue (Fig. 5G) but was clearly absent in newly created invasive strands (Fig. 5H, arrow). A transverse section showed not only strong LOX staining in the epithelial lining (Fig. 5I, J) but also in the stellate reticulum when evaluated at higher magnification (Fig. 5J, asterisk).

**Figure 4:**
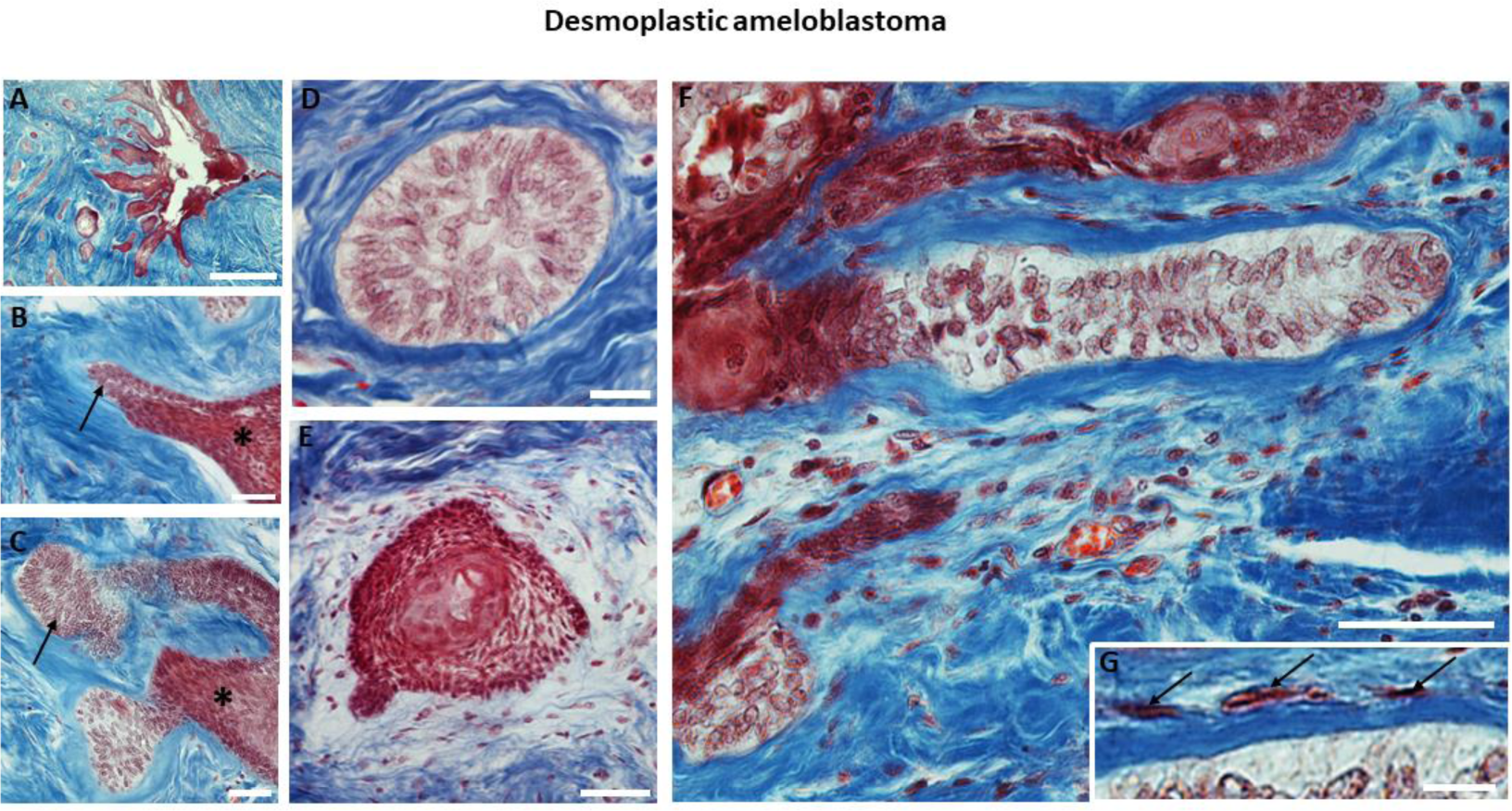
Tumour invasion and extracellular matrix remodeling in desmoplastic ameloblastoma using Masson’s Trichrome staining. A. Low magnification showing the stromal (blue) and tumorigenic tissue (red). Higher magnification reveals sharp (B) and isle-shaped (C) tumorigenic outgrowths invading the stromal tissue. Note the presence of translucent tumour cells within these outgrowths (B, C; arrows) which are distinctively different from the dark tumour cell phenotype of the tumorigenic mass (B, C; asterisks). Transverse sections of outgrowths (D, E) showing either translucent tumour cells engulfed by dense cross-linked collagen fibrils (D) or darker isles with ameloblast-like cells and stellate reticulum encircled by loose organized collagen (E). F. Longitudinal section of a tumorigenic outgrowth with a translucent tumour cell phenotype engulfed by highly cross-linked collagen. Mark the absence of a stellate reticulum (F) and the periodic appearance of CAF interspersed between the collagen fibrils (higher magnification of F in G; arrows). Scale bars represent 25 (G), 50 (D), 100 (B, C, E, F), 500 (A) μm.

**Figure 5:**
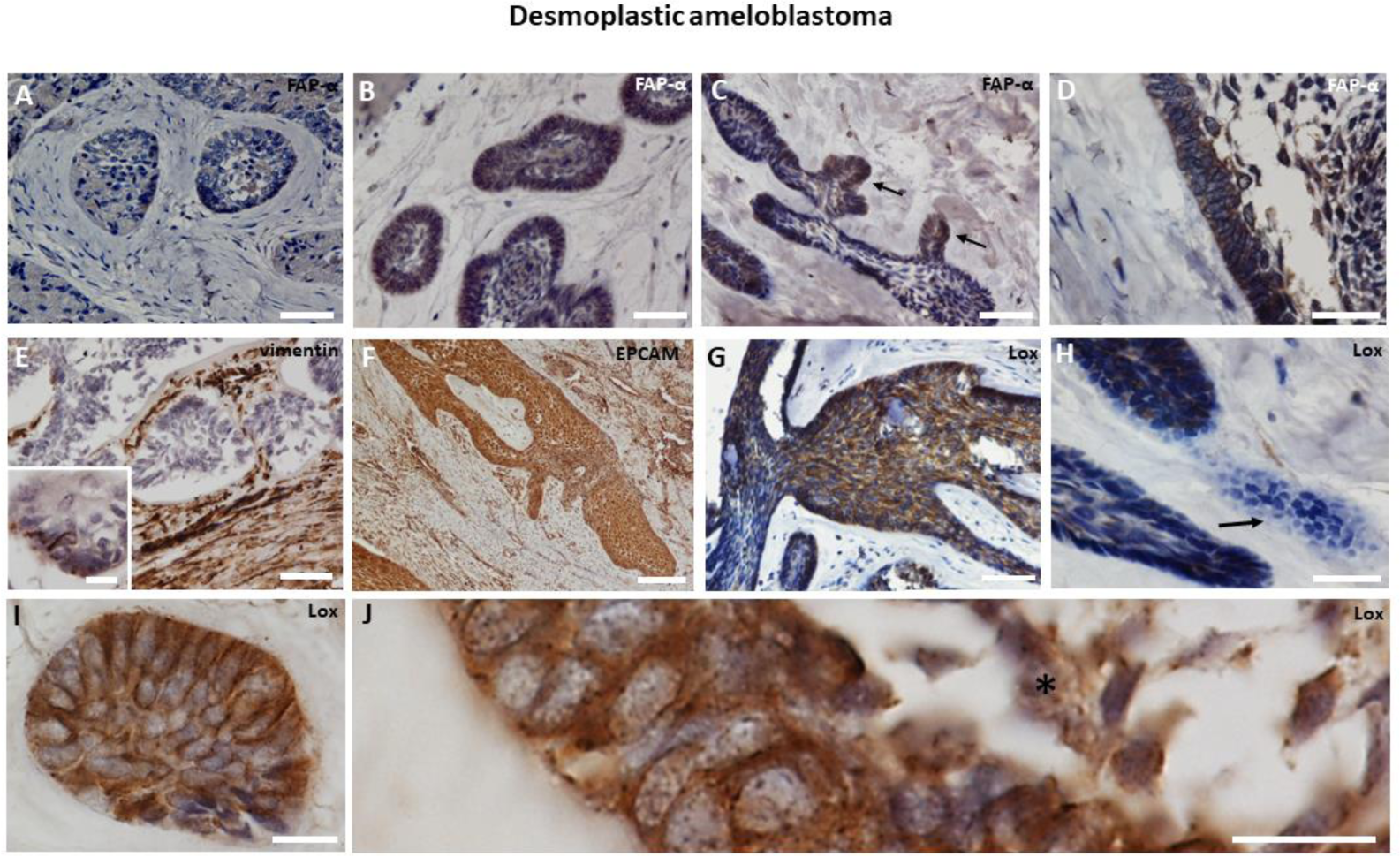
Identification of FAP-α and LOX expressing cells in desmoplastic ameloblastoma. A. Immunohistochemistry showing weak FAP-α staining in tumorigenic isles surrounded by dense connective tissue. Strong FAP-α staining in tumorigenic islands (B) and protrusions (C, arrows) with ameloblast-like cells and stellate reticulum (D). E. Vimentin staining discriminates between the highly positive stroma and the negative tumorigenic tissue. Insert shows a few vimentin positive cells in the ameloblast-like cell region. Strong EPCAM (F) and LOX (G) staining observed in the tumorigenic tissue. H. Absent LOX staining in newly formed tumorigenic tissue (arrow). I-J. Higher magnification of I shows strong LOX staining in the ameloblast-like cells and stellate reticulum (J, *). Scale bars represent 25 (I, J, E insert), 50 (D, H), 100 (A, B, C, E, G) and 250 (F) μm.

### Quantification and comparison of FAP-α expression in solid ameloblastoma

To evaluate differential FAP-α expression within the tissue and between subtypes of solid ameloblastoma, we quantified FAP-α positive cells in respectively three regions of each subtype i.e. the epithelial lining, the stellate reticulum and the stromal tissue. In follicular ameloblastoma, FAP-α staining was noted in the majority of the ameloblast-like cells (51 ± 9% of the cells, Fig. 6A) but the total number of FAP-α positive cells was significantly lower in the stellate reticulum (17 ± 5%; p < 0.05) and the matching stromal tissue (12 ± 3%; p < 0.01). In plexiform ameloblastoma, a high number of FAP-α positive cells was present in the ameloblast-like epithelial layer (Fig. 6A; 62 ± 12%) but was not significantly different from the stellate reticulum and stromal cells. In desmoplastic ameloblastoma, a high concentration of FAP-α positive ameloblast-like cells was detected in the epithelial lining (57 ± 9%; Fig. 5D, Fig. 6A) but the number of FAP-α positive cells was markedly lower in the stellate reticulum (25 ± 4%; Fig. 6A) and stroma (20 ± 3%; Fig. 6A). Consequently, we compared the number of FAP-α positive cells specific for each region between the three subtypes (Fig. 6B). The concentration of FAP-α positive cells in the epithelial border and stroma was comparable between subtypes. However, the stellate reticulum of plexiform ameloblastoma contained a significantly higher concentration of FAP-α positive cells than the corresponding region in follicular ameloblastoma (Fig. 6B, 50 ± 14% *vs* 17 ± 5% respectively; p < 0.05).

**Figure 6:**
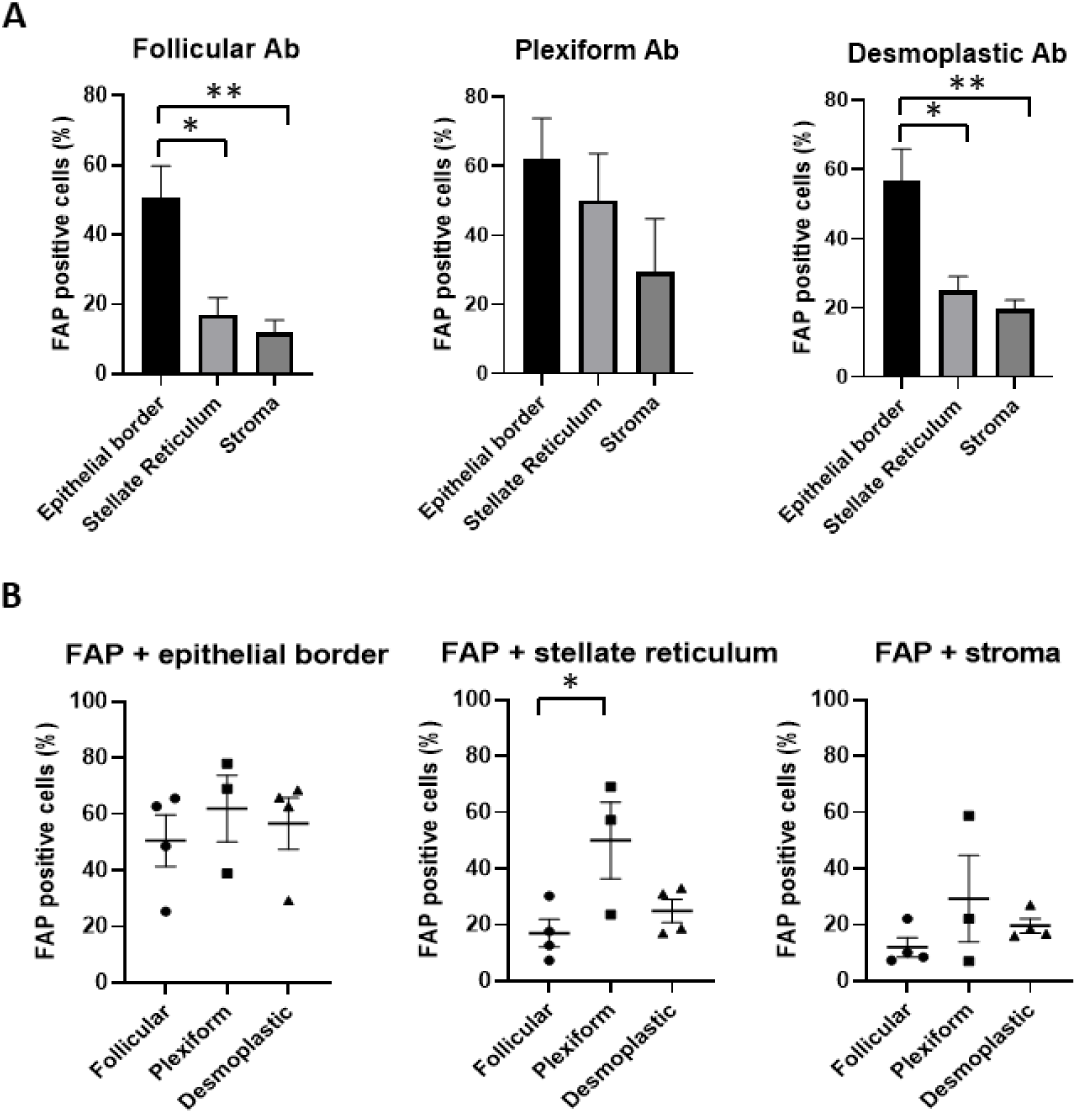
Quantification of FAP-α expressing cells in subtypes of solid ameloblastoma within the connective and tumorigenic tissue. A. Comparison of the percentage of FAP-α positive cells in the epithelial border, stellate reticulum and stroma within the follicular (n=4), plexiform (n=3) and desmoplastic ameloblastoma (n=4). B. Comparison of the percentage of FAP-α positive cells between regions of different ameloblastoma subtypes. (* p < 0.01; ** p < 0.05)

### FAP-α expression in unicystic ameloblastoma

Finally, we studied FAP-α and LOX expression in three distinct types of unicystic ameloblastoma. Type I displayed a thin odontogenic epithelium positive for FAP-α (Fig. 7A) and LOX (Fig. 7B, C) whereas type II showed a FAP-α (Fig. 7D) and LOX (Fig. 7E) positive epithelium with intraluminal tumorigenic cells. Type III was characterized by FAP-α positive tumorigenic cells with features of plexiform ameloblastoma (Fig. 7F). Ameloblast-like cells stained highly positive as well as the stellate reticulum (Fig. 7G) with the presence of FAP-α negative giant cells (arrow). The distribution of tumorigenic tissue was identified by LOX staining (Fig. 7H) and showed a similar staining pattern as FAP-α when visualized at higher magnification (Fig. 7I). Quantitatively, we determined that FAP-α expression was highly pronounced throughout all distinct regions of the unicystic subtypes including the ameloblast-like cells (70 ± 12%; n=3) and stromal tissue (40 ± 24%; n=3). The stellate reticulum which is only a feature of type III revealed a strong FAP-α staining in 49% (n=1) of the tumour cells.

**Figure 7:**
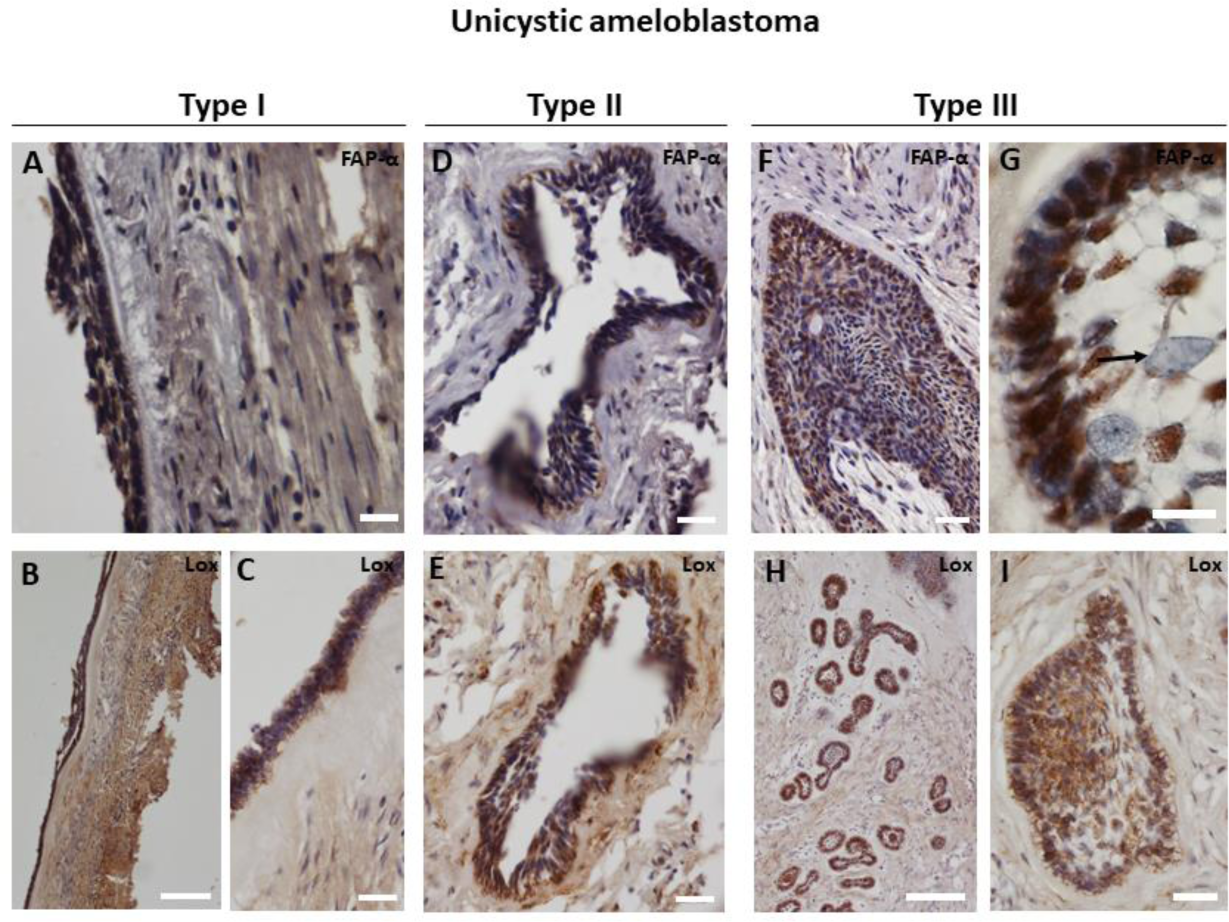
FAP-α and LOX expression in different types of unicystic ameloblastoma. Immunohistochemistry of thin odontogenic epithelium in type I showing FAP-α (A) and LOX (B, higher magnification in C) positive staining. Type II reveals a FAP-α (D) and LOX (E) positive epithelium with intraluminal proliferation of tumorigenic cells. FAP-α positive plexiform-like tissue in type III (F) with ameloblast-like cells and stellate reticulum (Fig. 7G). Note the presence of FAP-α negative giant cells (G, arrow). Identification of type III tumorigenic tissue by LOX staining at low (H) and high magnification (I). Scale bars represent 25(G), 50 (A, C, D, E, F, I), 250 (B, H) μm.

## Discussion

In this study, we show that FAP-α is highly expressed within the tumorigenic tissue of ameloblastomas and in particular in the plexiform and type III unicystic ameloblastoma. Our study reveals differences in regional LOX expression between subtypes of ameloblastoma which suggest a differential mechanism of tumour growth and ECM remodelling. Tumour invasion and growth in ameloblastoma tissue has been associated with matrix metalloproteinases expression ^24^. These enzymes actively degrade the basal membrane and ECM allowing directional growth of tumorigenic cells. MMP-9 displays the highest expression in plexiform ameloblastoma compared to follicular ameloblastoma coinciding with a higher invasive capacity ^25,26^. Similarly, plexiform ameloblastoma denoted in our study a significant higher concentration of FAP-α positive cells in the stellate reticulum compared to follicular ameloblastoma. Since collagen degrading enzymes are abundantly present, we hypothesize that these enzymes contribute to the tumour invasive capacity and growth. However, whether this reflects a higher tumour growth rate in plexiform ameloblastoma compared to follicular ameloblastoma remains to be investigated. A common observation in all ameloblastoma subtypes is the high concentration of FAP-α positive cells in the epithelial border of the tumorigenic tissue. Similar findings are reported for MMP-2 ^27^ and podoplanin ^28^ expression. These studies support the hypothesis that tumorigenic invasion is regulated through collagenase activity at the epithelial ameloblast-like cell lining. A recent report suggests that local invasiveness of ameloblastomas is determined by EMT activation regulated via TGF-β1 ^29^ and thus would enhance the migratory capacity of tumour cells into the connective tissue. Since FAP-α expression is considered an EMT marker ^19^ and is highly expressed in ameloblastomas, it would imply that EMT is the most prominent mechanism for tumour growth. However, in our study FAP-α positive tumorigenic tissue displays mainly epithelial characteristics originating from the initial odontogenic epithelium as shown by high EPCAM expression and absence of vimentin expression. A sub-population of epithelial tumour cells from breast and prostatic cancer have been described to co-express EPCAM and FAP-α indicating that FAP-α is not solely and specifically expressed by CAF ^30^. Vimentin positive cells are only sporadically observed inside the tumorigenic tissue of our samples. This is in contrast to epithelial carcinomas in which high vimentin expression is correlated to the grade of malignancy and metastasis ^31,32^. We hypothesize that the benign ameloblastoma tissue is in a state of intermediary EMT granting the tissue the capacity to produce collagenases for local invasion but without the ability to metastasize. We should not rule out that single EMT-converted epithelial tumour cells detach from the tumour tissue and migrate into the stroma but these are likely to be confined to the mesenchymal cell pool.

Local micro-environmental changes which enhance tumour invasiveness are not only determined by connective tissue degradation but also by creating a stiffer ECM. A previous study in carcinoma revealed that the ECM was remarkably stiffer at the invasion front of the growing tumour ^22^. ECM stiffness in tumours is regulated by CAF which express and secrete LOX, a putative cross-linker of collagen type I. A stiffer matrix creates a positive feed-back loop leading to a higher CAF number, activation of the TGF-β1 pathway, FAP-α expression and ultimately a higher LOX production ^20^. This mechanism would certainly fit desmoplastic ameloblastoma due to the high content of connective tissue, TGF-β1 ^33^ and the FAP-α positive CAFs described in our study. However, LOX staining is only prominent in tumorigenic tissue of desmoplastic and unicystic ameloblastoma and not within stroma suggesting a differential mechanism when compared to carcinoma. Both desmoplastic and unicystic ameloblastoma are mainly composed of connective tissue which could impose high mechanical load on tumour cells and promote LOX expression via focal adhesion kinase (FAK) phosphorylation. FAK activation has been correlated with increased ECM stiffness and high expression was found in ameloblastoma ^34^. High LOX expression in tumorigenic tissue can aid in cross-linking of the adjacent ECM or as reported by others stimulate tumour cell invasion and proliferation ^20^. We report that tumorigenic outgrowths are accompanied by aligned spindle-shaped cells resembling CAF which can assist in guiding tumour invasion as reported in carcinoma. Indeed, a reciprocal interaction between fibroblasts and the AM-3 ameloblastoma cell line has been proven in 3D cell culture in which interleukin secretion by fibroblasts enhanced tumour invasion and migration ^11^. Based on our histological observations, we suggest a multi-step mechanism in desmoplastic ameloblastoma invasion: 1) CAF recruitment and LOX-induced collagen cross-linking at the tumour invasion front, 2) ECM degradation by FAP-α, 3) phenotypic switch into a FAP-α and LOX negative tumorigenic cell type followed by migration and proliferation, and 4) differentiation of newly formed tumorigenic cells into FAP-α and LOX positive ameloblast-like cells and stellate reticulum.

When evaluating LOX expression in solid ameloblastoma subtypes, follicular and plexiform ameloblastoma are markedly different from desmoplastic ameloblastoma. LOX staining in follicular ameloblastoma is also present in the tumorigenic tissue but rather weak whereas the plexiform ameloblastoma reveals an abundant staining pattern restricted to the stroma. From these observations, we suggest that plexiform ameloblastoma shares a similar LOX staining pattern as seen in epithelial carcinoma which can be related to the aforementioned positive feedback-loop mechanism regarding matrix stiffness and invasion. The weak or even absent LOX expression in follicular and plexiform ameloblastoma can be explained by the higher ratio of tumorigenic tissue to connective tissue resulting in a lower mechanical load imposed on tumour cells. In conclusion, we suggest that FAP-α and LOX expression are major contributors to ameloblastoma growth and ECM remodelling. Regional LOX expression is differentially regulated and specific for each ameloblastoma subtype which offers a new tool for understanding the growth patterns characteristic for each subtype.

### Future perspectives

Targeting FAP-α expression is a promising approach in reducing tumour growth and invasion in carcinoma ^10^. Several specific blockers have been shown to procure a significant effect in arresting carcinoma development. In addition, inhibition of LOX expression or activity has been shown to suppress tumour growth by interfering in the MATN2 expression and EGFR pathway ^35^. The results from this study could offer alternative approaches in arresting ameloblastoma growth and in prevention of recurrent growth by targeting LOX and FAP-α or by inhibiting the downstream signalling pathway of both enzymes.

## Data Availability

All data produced in the present study are available upon reasonable request to the authors.

## Acknowledgements

We would like to thank Marc Jans and Jeanine Santermans (Hasselt University) for the histological preparation of the samples.

## Author Contributions

TV and RBD conceived, designed, performed the histological staining, analysed and interpreted the data. RBD and TV wrote the paper which was approved by all co-authors. PG, EW and IL conceived, designed, interpreted the data and reviewed the paper. JOA, AOA, AAO, JTA, CP conceived and reviewed the paper.

